# Radio-pathomic maps of histo-morphometric features trained with whole mount prostate histology distinguish prostate cancer on MP-MRI

**DOI:** 10.64898/2026.01.09.26343808

**Authors:** Savannah R. Duenweg, Samuel A. Bobholz, Allison K. Lowman, Aleksandra Winiarz, Biprojit Nath, Michael J. Barrett, Fitzgerald Kyereme, Stephanie Vincent-Sheldon, Kathleen Bhatt, Katherine Troy, Michael Kim, Eric Fair, Kenneth A. Iczkowski, Kenneth M. Jacobsohn, Anjishnu Banerjee, William A. Hall, Andrew S. Nencka, Peter S. LaViolette

## Abstract

**Background:** Prostate cancer (PCa) is the most prevalent male cancer in the U.S., accounting for 29% of new cancer diagnoses. Multiparametric MRI (MP-MRI), including T2-weighted imaging (T2WI) and apparent diffusion coefficient (ADC) maps, is an effective tool for detecting PCa; however, accuracy varies, and false-positives may lead to unnecessary biopsies or overtreatment. Radio-pathomic maps (RPMs), derived from MP-MRI and machine learning, have been advantageous in differentiating clinically significant PCa. This study tested whether RPMs of tissue density and histo-morphometric features could better predict cancer presence than conventional MR imaging.

**Materials and Methods:** MP-MRI from 236 patients prospectively recruited between 2014 and 2023 with confirmed PCa were analyzed. Whole-mount prostate sections sliced to match the MRI were processed, digitized, and Gleason-pattern annotated by a GU pathologist. Automated algorithms identified glands and calculated quantitative histo-morphometric features, which were mapped across whole slide images. Slides were nonlinearly aligned to each patient’s T2WI using in-house software, enabling direct comparison of slides, features, and annotations in MR-space. A multi-step prediction model was trained using a 2/3 – 1/3 train/test split to predict histo-morphometric features using 5×5 voxel tiles from T2WI and ADC. These feature maps were then used generate tumor probability maps.

**Results:** Histological feature models produced RMSE values approximately within one standard deviation of the ground truth’s variability, indicating acceptable performance. The best RPM, using histological density features, achieved an accuracy of ∼80%. Visual inspection of RPMs showed good concordance to high-grade cancer annotations.

**Conclusion:** This study demonstrates that the use of MRI intensities can predict complex histo-morphometric features and delineate regions of PCa non-invasively. Future research is warranted to determine the clinical benefit of using RPMs in treatment guidance.

## 1. Introduction

Prostate cancer (PCa) is a leading male cancer worldwide, with a 3% annual incidence rate rise in the United States. In the U.S., PCa is projected to be responsible for over 35,000 cancer-related deaths in 2024(1), making it a significant health concern. While highly prevalent, not all cases pose a high risk of metastasis or mortality(2, 3). Understanding the spectrum of PCa’s severity and individual risk factors is imperative for effective management and treatment.

Multiparametric magnetic resonance imaging (MP-MRI) has emerged as a valuable technique in the evaluation of PCa, enabling more accurate diagnoses(4, 5). The implementation of the Prostate Imaging Reporting and Data System (PI-RADS) has significantly contributed to the standardization of prostate MRI practices, encompassing acquisition, interpretation, and reporting protocols. PI-RADS provides scores between 1 and 5 to lesions detected on T2-weighted images (T2WI) and diffusion weighted images (DWI) with corresponding apparent diffusion coefficient (ADC) maps(6). By providing a structured framework, PI-RADS has enhanced the detection of cancerous lesions and fostered greater consistency in radiology assessments, although there are still limitations to PI-RADS scoring, particularly in small tumors or equivocal cases(7, 8).

While MP-MRI has aided in lesion detection, the underlying pathology remains unknown until biopsy or surgery. Prostate histology is graded using the Gleason grading scale, assigning a score from the grades of the two most dominant glandular patterns. Recently, Gleason grades have been condensed into one of five Grade Groups (GG) to predict patient prognosis(9). Low risk cancers may be managed through active surveillance, where patients receive regular prostate specific antigen (PSA) blood tests. If clinically significant cancer is found at biopsy or following an increased PSA test, patients may be treated with surgery and/or other therapies. Although PI-RADS and GGs are the gold standard practices for detecting and diagnosing PCa, these are both qualitative assessments, and thus risk interobserver variability(8, 10).

Under-diagnosing PCa may result in cancer development; overtreatment may result in unnecessary biopsies and surgeries, which themselves may lead to infection, incontinence, and impotence(11). Radio-pathomic mapping is an innovative technique that leverages datasets of aligned MRI and digital pathology to train machine learning models(12). Correlating radiological and histological features can uncover valuable insights into tumor presence, heterogeneity, and severity(13, 14) non-invasively. This study used whole mount digitized PCa histology with pathological annotations to determine imaging signatures predictive of PCa presence. Specifically, we tested the hypothesis that MP-MRI intensity values are predictive of pathomic features of PCa and that a radio-pathomic model trained on this data can accurately delineate regions of PCa beyond current imaging guidelines.

## 2. Materials and Methods

### 2.1. Patient Population and Data Acquisition

This institutional review board-approved retrospective study analyzed data from 432 prospectively recruited patients with biopsy-confirmed PCa undergoing radical prostatectomy between 2014 and 2023. Written, informed consent was obtained from all patients for study participation. Inclusion criteria included having clinical T2WI and ADC maps, as well as pathologist-provided annotations (see Section 2.3). A total of 236 patients (mean age 61.9 ± 6.6 years) met these criteria and were separated utilizing a 2/3 – 1/3 train/test split for model development, to ensure model generalizability, balanced by Gleason Grade Group. Detailed clinicopathological characteristics of the patient cohort are presented in **Table 1**.

**Table 1.** Clinicopathological features of the prostate cancer cohort assessed at time of surgery. Race was self-reported upon recruitment and not further analyzed in this study. *Abbreviations: RP = radical prostatectomy, PSA = prostate specific antigen*

|  | Training<br>(n = 158) | Testing<br>(n = 78) | Total<br>(n = 236) |
| --- | --- | --- | --- |
| Age at RP, years (mean, SD) | 61.6 (6.3) | 62.4 (7.2) | 61.9 (6.6) |
| Race (n, %) |  |  |  |
| African American | 13 (8.2) | 11 (14.1) | 24 (10.2) |
| White/Caucasian | 144 (91.1) | 65 (83.3) | 209 (88.6) |

| Other* | 1 (0.6) | 2 (2.6) | 3 (1.3) |
| --- | --- | --- | --- |
| Preoperative PSA, ng/mL (n, %) |  |  |  |
| < 6 | 75 (47.5) | 33 (42.3) | 108 (45.8) |
| ≥ 6 – 10 | 55 (34.8) | 28 (35.9) | 83 (35.2) |
| ≥ 10 – 20 | 26 (16.5) | 14 (17.9) | 40 (16.9) |
| ≥ 20 – 30 | 2 (1.3) | 3 (3.8) | 5 (2.1) |
| Grade group at RP (n, %) |  |  |  |
| 6 | 16 (10.1) | 8 (10.3) | 24 (10.2) |
| 3+4 | 95 (60.1) | 46 (59.0) | 141 (59.7) |
| 4+3 | 24 (15.2) | 12 (15.4) | 36 (15.3) |
| 8 | 4 (2.5) | 2 (2.6) | 6 (2.5) |
| ≥ 9 | 19 (12) | 10 (12.8) | 29 (12.3) |
| Clinical Stage (n, %) |  |  |  |
| T1 | 132 (83.5) | 62 (79.5) | 194 (82.2) |
| T2 | 26 (16.5) | 16 (20.5) | 42 (17.8) |
| Surgical Stage (n, %) |  |  |  |
| 2a,b | 85 (53.8) | 40 (51.3) | 125 (53) |
| 2c | 23 (14.6) | 12 (15.4) | 35 (14.8) |
| 3a,b | 50 (31.6) | 26 (33.3) | 76 (32.2) |

\* Other = Asian, Hispanic, or self-reported “Other”

### 2.2. MR Image Acquisition and Pre-Processing

All patients underwent clinical MP-MRI on a 3T scanner prior to radical prostatectomy. Imaging protocols included T2WI, and field-of-view (FOV) optimized and constrained undistorted single shot (FOCUS) DWI with a range of b-values. As the images that we used in this study were all clinically obtained, there were a variety of MRI acquisition parameters including manufacturer (General Electric, Waukesha, WI, USA; Siemens Healthineers, Erlangen, Germany; Toshiba, Minato, Tokyo, Japan; or Philips Healthcare, Amsterdam, Netherlands; n = 163, 65, 4, and 4, respectively); slice thickness (3 mm = 211; 4 mm = 19, 3.2 mm = 1; 3.5 mm = 1; 2.5 mm = 4); average repetition and echo time = 4417.5 and 112.2 ms, respectively; coil (endorectal coil = 62, scanner specific 32Ch body coil = 174). ADC maps that were not generated straight from the MR scanner were calculated between b-values of 0-1000 due to the inconsistency in imaging collected across patients. T2WI underwent z-score intensity normalization using a mask of the whole prostate. ADC maps were aligned to the T2WI using ITK Snap’s *tkregister2* (www.itksnap.org) and manually adjusted as necessary. We opted to use T2WI and ADC exclusively in this study as they were most frequently available across all patients. A recent study found that bi-parametric MRI performs as well as MP-MRI at prostate cancer detection, supporting our decision to limit our analysis to these two sequences(7).

### 2.3. Histological Processing

Robotic prostatectomy was performed approximately 8 weeks following imaging using the da Vinci system (KMJ, >20 years of experience) (Intuitive Surgical, Sunnyvale, CA, USA)(15, 16). Prostate samples were formalin-fixed overnight and sectioned using patient-specific tissue slicing jigs that enable direct one-to-one comparison between MRI and pathology using a previously published methodology (17). Briefly, prostate masks were manually drawn from patients’ T2WI using AFNI (Analysis of Functional NeuroImages, http://afni.nimh.nih.gov/)(18), modeled using 3dSlicer (slicer.org), imported into Blender 2.75 (https://www.blender.org/), and 3D printed using a fifth-generation Makerbot (Makerbot Industries, Brooklyn, NY, USA), creating a slicing jig to match the orientation and slice thickness of each patient’s T2WI(19, 20). A graphical representation of this process is shown in **Figure 2**.

**Figure 2.**
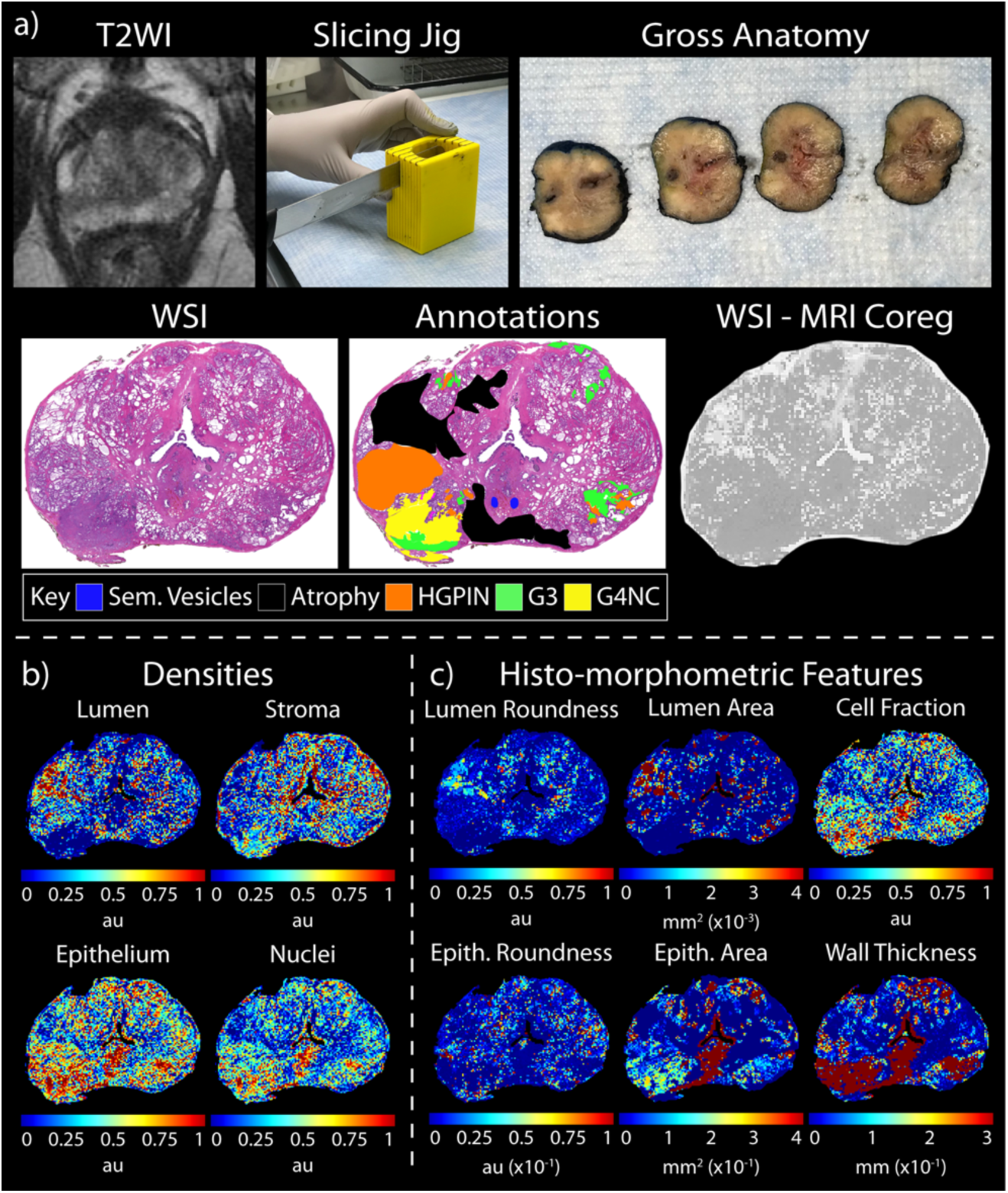
Breakdown of histological and radiological processing. a, top) Tissue is sectioned in line with patient’s T2WI, H&E stained, and digitized. a, bottom) Digitized slides are processed, annotated, and b) histological densities and c) histo-morphometric features are calculated. Finally, slides, annotations, and features are co-registered to the corresponding slice of the T2WI (a, bottom right).

Whole-mount tissue sections were hematoxylin and eosin (H&E) stained and digitized at 40x magnification using a sliding stage microscope (Huron Digital Pathology, Ontario, Canada) at a resolution of 0.2 μm/px. Due to the large file size, a downsample factor of 10 was employed for faster image processing. A total of 708 slide (n = 3/patient), chosen based on tissue quality and cancer presence, were manually Gleason pattern annotated by a GU-fellowship trained pathologist (KAI, >20 years of experience) using a Microsoft Surface Pro 4 (Microsoft, Seattle, WA, USA) or OMERO (Open Microscopy Environment, https://www.openmicroscopy.org), a secure end-to-end digital pathology platform (Figure 1), enabling us to have the most accurate ground truth label of the underlying pathology(21). The switch to OMERO allowed the storage of high-resolution images and shareability through a VPN connection, creating a convenient platform for annotations. Annotations from 150 slides were performed on whole slide images (WSI) generated on one of two lower resolution slide scanners at 40x magnification and resolutions of 0.85 μm/px (Nikon Metrology, Tokyo, Japan) (n = 114 slides) or 0.34 μm/px (Olympus Corporation, Tokyo, Japan) (n = 36 slides) prior to equipment upgrades. These slides were re-scanned, and annotations were brought into the higher resolution space of the current slide scanner. Annotation classes include seminal vesicles, atrophy, high-grade prostatic intraepithelial neoplasia (HGPIN), Gleason 3 (G3), Gleason 4 non-cribriform glands (G4NC), Gleason 4 cribriform (to papillary) glands (G4CG), and Gleason 5 (G5). G4CG and G4NC were separately annotated as significant prognostic disparities exist between the two(22). Seminal vesicles, atrophy, HGPIN, and non-annotated tissue were all classified as non-prostate cancer (nPCa) for simplified model training. G3 was additionally classified as nPCa as it is the divide between watchful waiting and aggressive treatment(23). All G4+ patterns were grouped as one cancer class.

### 2.4. Histo-morphometric Feature Calculation

To expedite processing time, WSI were downsampled by factor of 2(24) and custom, in-house Matlab (Mathworks Inc, Natick, MA) pipelines were employed to extract pathological features for quantitative analysis following a previously published method(24, 25). Briefly, a color deconvolution algorithm was implemented to segment stroma, epithelium, epithelial nuclei, and lumen based on their corresponding stain optical densities (i.e., positive hematoxylin or eosin and background). Next, features underwent filtering and smoothing to eliminate noise and enhance segmentations. These resulting masks were used to create density feature maps (DFM) of lumen, stroma, epithelium, and epithelial nuclei (LDS, SDS, EDS, CDS, respectively). Additionally, gland-level histo-morphometric features (HMFM) were computed including lumen roundness and area (LR, LA); epithelial roundness, area, and wall thickness (ER, EA, ET); and cell fraction (i.e., the ratio of epithelial cells per total gland area, defined by the area of the epithelial wall) (CF).

Epithelial wall thickness was demarcated as the shortest distance between the inner and outer gland boundaries. Examples of the pathomic feature calculations across a WSI are shown in Figure 2.

### 2.5. Histology & MRI Co-Registration

Digitized WSI, as well as their respective annotations and feature maps, were co-registered to the T2WI using established software and techniques(17, 26) to enable comparison between histology and clinical imaging through a local weighted-means transform, effectively accommodating non-uniform distortions. Briefly, slides were adjusted for flipping and rotation to match their respective MRI slice, and a control-point co-registration method was employed, manually placing 20-30 points on both modalities, positioned along the boundaries and distinctive landmarks within the prostate (i.e., urethra, seminal vesicles, etc.). A nonlinear spatial transformation was computed from these control points to downsample WSI to match MRI resolution. An additional nearest-neighbor interpolation was applied to maintain integer values to annotations. Finally, regions of interest were drawn excluding areas of tissue distortion (i.e., rips, tears, and folds) and MRI artifacts (Figure 2).

### 2.6. Histological Feature Analyses

Linear mixed-effects models (LMMs) were used to determine differences between calculated DFM and HMFM within regions of cancer and non-cancer. Features were averaged within lesions and non-cancerous tissue across each slide. Cancer status per lesion was included as a main effect, with patient, slide, and slide nested within patient as random effects to account for these confounding factors. Parameters of estimates and p-values were reported to quantify the relationship between each feature and PCa presence.

### 2.7. Radio-pathomic Maps of Prostate Cancer using Histo-morphometric Features and MRI Intensities

Creation of prostate radio-pathomic maps (RPMs) involves a multi-step machine learning model following a similar architecture used in glioblastoma imaging(27). All features were normalized using the z-score of the training data. Bootstrap aggregating (bagging) random forest regression models were trained to predict DFM and HMFM using 5×5 voxel tiles from the T2WI and ADC as input. Root-mean-squared error (RMSE) estimates were used to quantitatively evaluate performance in the held-out test set, and example maps were generated to qualitatively compare to ground truth segmentations (Figures 3 **and 4**).

**Figure 3.**
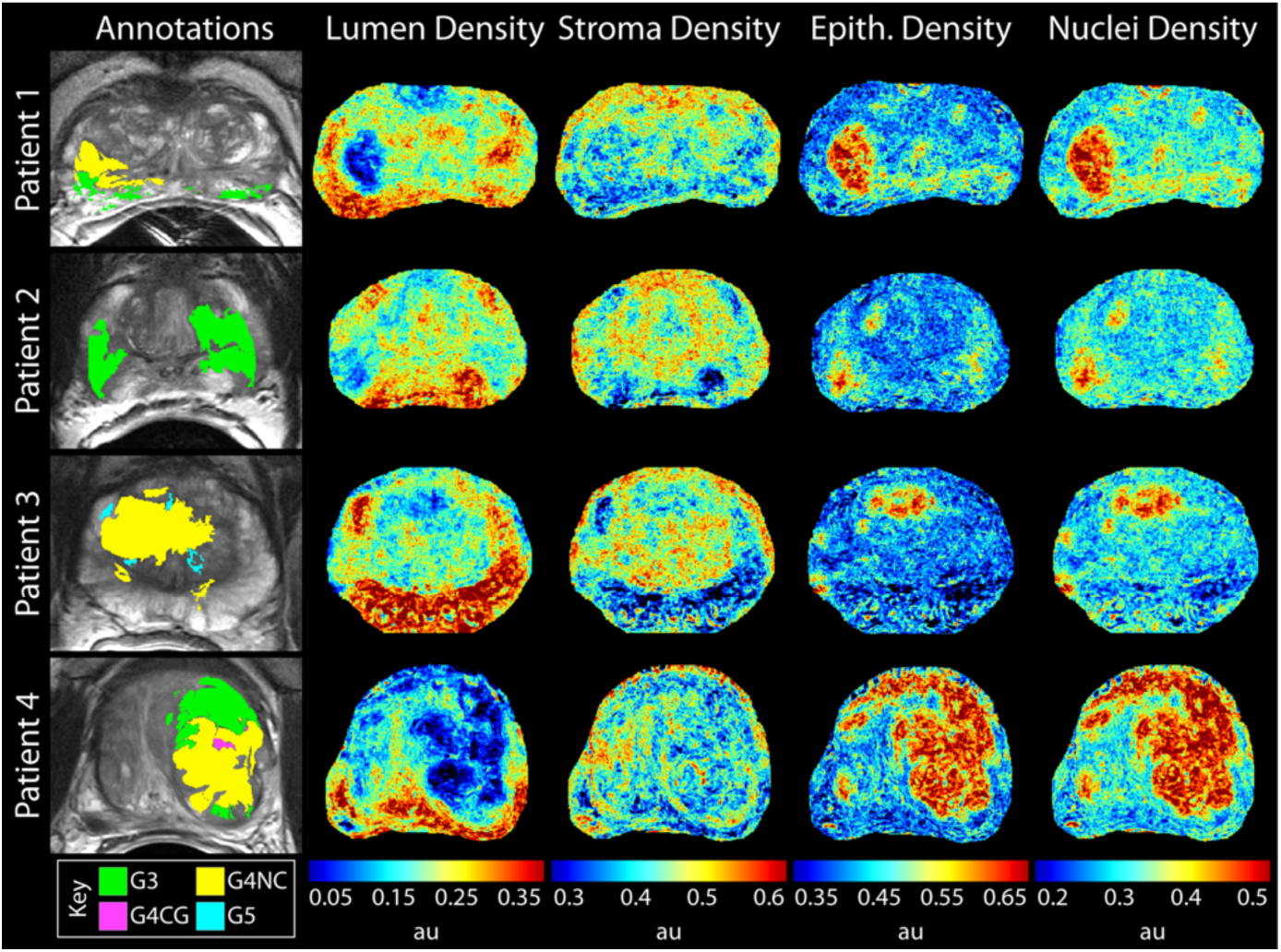
Tissue component density maps compared to ground truth annotations (left). Lumen density was found to be low (second column), and epithelium and cell densities (right columns) were high in regions of high-grade cancer.

**Figure 4.**
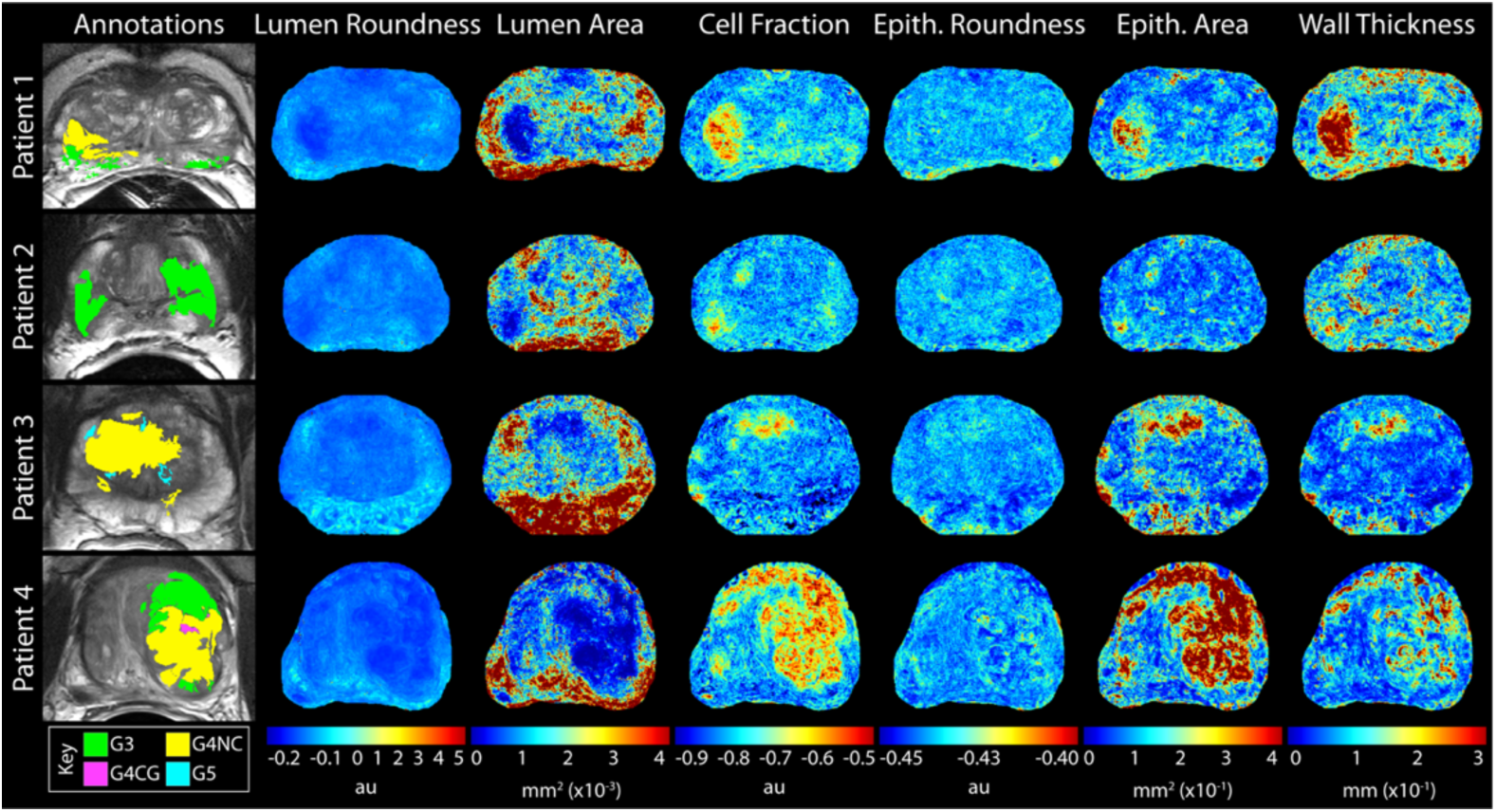
Second order feature maps compared to ground truth annotations (left). Luminal features were low in regions of tumor and epithelial features were high in regions of high-grade cancer.

Next, the ground truth segmentations of each feature were used as input to RUSBoosted ensemble classification models with a 5-fold cross validation to predict voxel-wise cancer presence. These models were trained using DFM, HMFM, and all features (AFM), for a total of 13 individual histology-based models (10 histology regressions, 3 classifiers). RPM model performance was evaluated on the test set and RPMs were created using the predicted histological feature maps and compared to ground truth pathological annotations (Figure 5). Predicted features and RPM values were compared using LMMs with the same modeling parameters as previously describd to determine if 1) RPMs captures the same relationship between features and cancer, and 2) RPMs differ in cancerous regions.

**Figure 5.**
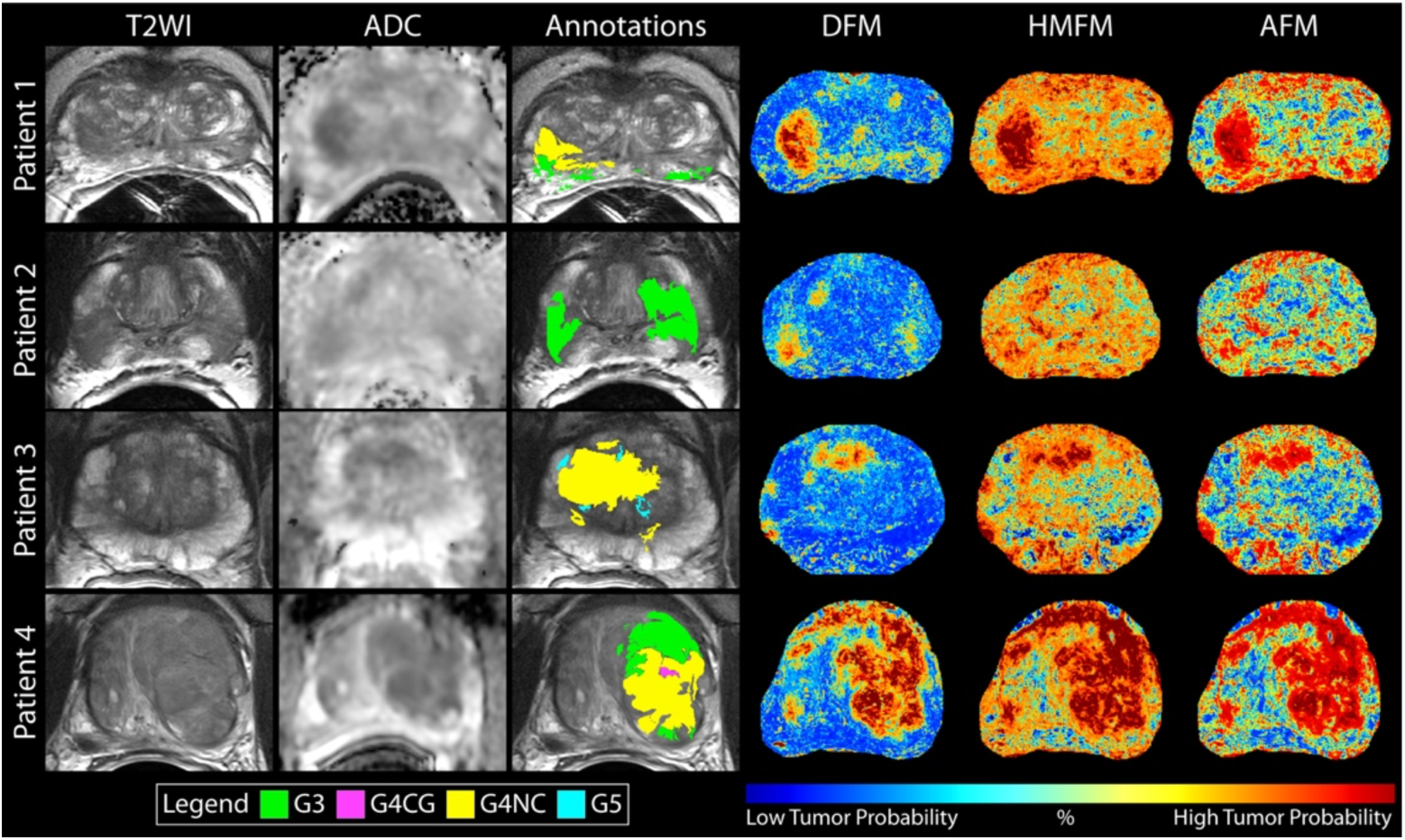
Clinical T2WI and ADC maps compared to pathological annotations (middle) and radio-pathomic maps of tumor probability (left). Density feature maps (DFM) best delineate regions of suspected tumor compared to histo-morphometric feature (HMFM) and the all feature (AFM) maps.

## 3. Results

### 3.1. Histological Feature Analyses

Mixed-effects model results for the histological feature analyses are presented in **Table 2 (left)**, where the reference group was non-cancer. LDS, SDS, LR, and LA were all higher in non-cancerous regions (LDS β = 46.51, p < 0.001; SDS β = 5.99, p = 0.03; LR β = 0.05, p < 0.001; LA β = 0.001, p < 0.001). Conversely, EDS, CDS, EA, ET, and CF were all lower in non-cancerous regions (EDS β = −52.62, p < 0.001; CDS β = −55.71, p < 0.001; EA β = −0.18, p < 0.001; ET β = −7.68E-4, p < 0.001; CF β = −0.16, p < 0.001). ER was not statistically different between groups; however, it trends towards being lower in non-cancerous regions (β = −0.02, p = 0.06).

**Table 2.**
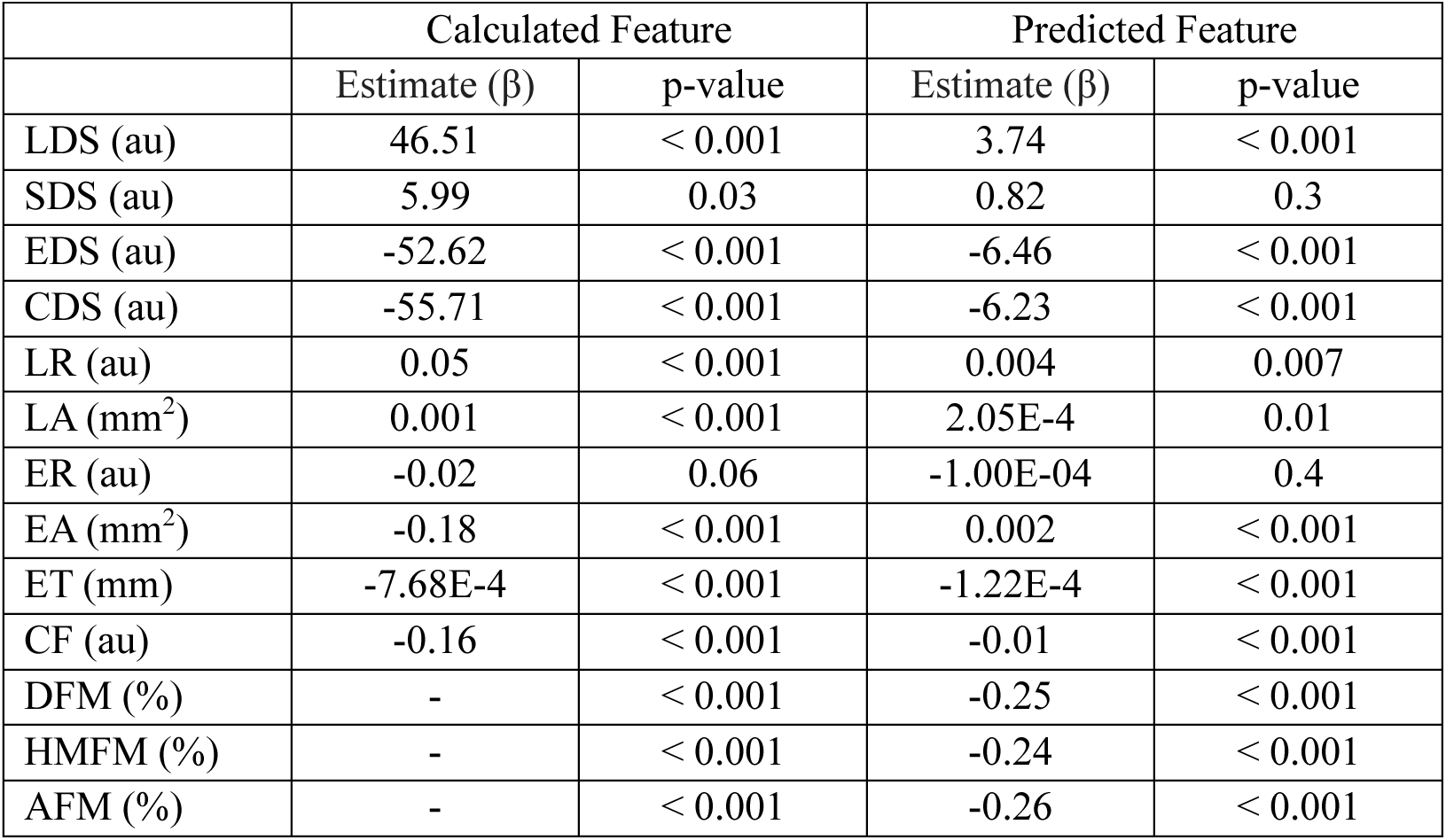
Calculated and predicted feature results from linear mixed models. Units for each feature is presented on the left column with their respective names.

Predicted features primarily followed these same trends, with LDS, LR, LA predicted larger in non-cancerous lesions (LDS β = 3.74, p < 0.001; LR β = 0.004, p = 0.007; LA β = 2.05E-4, p < 0.001); however, EA predicted higher in non-cancer as well (β = 0.002, p < 0.001). EDS, CDS, ET, and CF were likewise lower in non-cancerous tissue regions (EDS β = −6.46, p < 0.001; CDS β = −6.23, p < 0.001; ET β = −1.22E-4, p < 0.001; CF β = −0.01, p < 0.001). Though predicted ER was also not statistically different between groups, it followed the same trend as the calculated features (β = −1.00E-4, p = 0.4).

### 3.2. Histo-morphometric Feature Prediction

Segmentation prediction results and examples from test set patients are shown in Figures 3 **and 4** for densities and histo-morphometric features, respectively. Lumen, stroma, and epithelium densities, as well as epithelial roundness, size, and wall thickness predictions exhibited a subject-level RMSE value that was at or under the ground truth feature’s standard deviation; the remaining four features had RMSE values < 1 above the standard deviation, indicating acceptable model performance for all tissue type (**Table 3**). Calculated and predicted lumen density was found to be low and epithelium and cell densities were high in regions of high grade tumors, as is consistent with previous studies(12).

**Table 3.** Histological segmentation prediction results. *Abbreviations: RMSE = root-mean squared error*.

| **Feature** | **RMSE** | **Standard Deviation** |
| --- | --- | --- |
| Lumen Density | 116.3 | 116.4 |
| Stromal Density | 128.3 | 129.5 |
| Epithelial Density | 116.3 | 130.5 |
| Epithelial Cell Density | 113.3 | 112.9 |
| Lumen Roundness | 0.56 | 0.55 |
| Lumen Area | 3.1e4 | 3.0e4 |
| Cell Fraction | 0.43 | 0.42 |
| Epithelial Roundness | 0.21 | 0.21 |
| Epithelial Size | 2.0e6 | 2.0e6 |
| Epithelial Thickness | 17.5 | 17.2 |

### 3.3. Prostate Radio-Pathomic Maps

The accuracy of the three radio-pathomic maps was analyzed using a left-out test dataset from 78 patients. All models performed at approximately 80% accuracy (DFM = 79.3%, HMFM = 81.5%, AFM 80.1%) (**Table 4**). The AFM had the greatest accuracy at predicting cancer at 72.3% compared to DFM which had the lowest per-class accuracy of 65.8%, however, DFM visually appear to have the best delineation between tumor and not tumor. HMFM have well defined margins of cancerous regions on the RPMs, however, they appear to edge towards higher cancer probability across the whole prostate. Notably, the DFM find regions of G3, considered noncancer in the training data, but at much smaller magnitudes than the histological annotations. Linear mixed model results determined nPCa regions were statistically lower than in cancerous lesions (β = −0.25, −0.24, −0.26, for DFM, HMFM, and AFM, respectively; all p < 0.001) (**Table 2**).

**Table 4.** Prostate radio-pathomic mapping models performances. Abbreviations: DFM = density feature maps; HMFM = histo-morphometric feature maps; AFM = all feature maps, nPCa = non-prostate cancer, PCa = prostate cancer

| Model | Accuracy<br>Overall [nPCa, PCa] | Precision<br>Overall [nPCa, PCa] | Recall<br>Overall [nPCa, PCa] | F1-Score |
| --- | --- | --- | --- | --- |
| **DFM** | 79.3 [82.8, 65.8] | 0.76 [0.80, 0.72] | 0.56 [0.99, 0.13] | 0.64 |
| **HMFm** | 81.5 [85.4, 65.9] | 0.77 [0.82, 0.72] | 0.56 [0.99, 0.14] | 0.65 |
| **AFM** | 80.1 [84.3, 72.3] | 0.77 [0.80, 0.74] | 0.56 [0.99, 0.14] | 0.65 |

## 4. Discussion

In this study, we developed an innovative, multi-stage tumor prediction model that identifies areas of prostate cancer beyond traditional imaging signatures delineated by PI-RADS. Our novel approach seamlessly integrates histological segmentations with MRI-based maps of glandular features, enabling a quantitative, non-invasive prediction of tumor presence with an accuracy of approximately 80%. This technique was validated using tissue samples collected following surgery and expertly Gleason patterns annotated, ensuring accuracy and reliability of our findings. These maps can provide clinical insights into the underlying tissue pathology, potentially aiding treatment decisions such as active surveillance, biopsy guidance, or targeted therapies.

While PI-RADS has significantly improved the detection of high grade PCa, there are notable limitations. There are vague concepts defined in the PI-RADS guidelines, specifically regarding PI-RADS 3 lesions, where it is visually harder to determine severity and thus treatment planning(28). PCa presence is defined by the size of the lesion and signal hypointensity, often described as a “smudged charcoal” appearance(29). While this helps differentiate benign or low-grade cancers from PI-RADS 4+, PI-RADS 3 can look like either high- or low-grade lesions. The use of DCE imaging is used in this case, however, its sensitivity to patient motion and low signal limit the ability to indicate severity(30). Additionally, this system is a qualitative assessment prone to inter-rater variability(31, 32), which has previously been shown to have low inter-rater agreement(33), highlighting the need for a quantitative assessment.

There has been a growing interest in studies predicting pathological grade and tumor habitats using MRI. A recent study in PCa radio-pathomic mapping used aligned T2, ADC, and WSI with annotations of aggressive and indolent lesions as input into a deep learning model that correlated features and then detected and characterized PCa lesions(34). Correlated feature learning extracted low-level textural characteristics between WSI and MR images which were used as input to a neural network, with an overall accuracy of 75% at detecting cancerous lesions and 73% at detecting clinically significant lesions. All three of our RPM models outperformed this model using interpretable, quantitative features derived from WSI; however, we did not explicitly determine the accuracy of our model predicting aggressive from indolent disease, as we labeled G3 as nPCa. An additional study using the Histo-MRI PCa cohort matched MR and histology images for use with machine learning(35). As with the first half of our RPM process, MRI is used as input for their algorithms which output histological features. This group likewise created molds using pre-operative imaging, however, it was used for *ex-vivo* imaging for correlation to a single slice of the prostate, which increases time and cost to the research group. These digitized slides were not aligned to the MRI, which may lead to result discrepancies if tissue warping occurs or slides left/right flipping or rotation occurred. Our work combats this by using tissue slicing molds created from *in-vivo* imaging and precisely aligning WSI to MRI for the best representation of MRI-histology findings.

### 4.1. Limitations

Although the findings of this study are promising, several limitations must be acknowledged. In comparision to previous studies, this study utilizes a relatively small patient cohort, thus employing a larger patient cohort may improve generalizability to external data. Additionally, this was a single-center study examining clinical imaging; therefore, lack of standardization in acquisition parameters may confound future studies using external data. Likewise, images both with and without an endorectal coil may create a bias to our analyses. Though the endorectal coil’s impact on MRI intensity is debated(36), it is known to alter the prostatic surface and may cause signal artifacts. Though our manually defined ROIs avoid signal artifacts, future studies are warranted to apply these models to a variety of MR images. Our registration methods likewise may cause artifacts due to the transformations that need to applied to either the ADC or histology; additionally these are both manual processes that are time consuming. Though we acknowledge this constraint on the registration process, no publically available method currently exists that consistently produces acceptable registrations on prostate MRI.

The use of multiple slide scanners of varying resolutions at the time of pathological annotation may be an additional confounding factor in our analyses. The decreased resolution of our previous slides may have hindered our pathologist’s ability to distinguish unique Gleason patterns. Similarly, annotations were only obtained from one pathologist and may not generalize to a larger populous of pathologists. Finally, ground truth for cancer presence was determined by our pathologist, which may not best inform a radiologist’s delination of cancer on MR imaging. Future studies should assess tumor presence from radiologist-annotated tumoral regions.

### 4.2. Conclusions

This study used digitized whole slide prostate cancer tissue samples from 236 patients aligned to MRI to develop a multi-stage model to predict cancer presence. This model accurately, non-invasively identified cancerous regions using quantitative histological features and MR imaging signatures. These results suggest that radio-pathomic mapping of PCa may provide non-invasive insight into cancer presence, which has the potential to aid in treatment decision making in real-world clinical cases. MRI intensity values were able to accurately predict histo-morphometric features at RMSEs within a standard deviation of the feature, and predictions from regressions were able to classify cancer presence with an overall accuracy of 80%. Future studies are warranted to determine if these features can delineate high-from low-grade cancer and unique Gleason patterns, as well as aiding in treatment planning including targeted biopsies.

## Data Availability

All data produced in the present study are available upon reasonable request to the authors

## Acknowledgments

We would like to thank our patients for their participation in this study.

## Notes

### Competing Interest Statement

The authors have declared no competing interest.

### Funding Statement

This research was funded by NIH/NCI R01CA218144, R21CA231892, R01CA249882, R01CA290631, Advancing a Healthier Wisconsin, Strain for the Brain Inc, the Ryan M. Schaller Foundation, and the State of Wisconsin Tax Check Off Program for Prostate Cancer Research.

### Author Declarations

The study was confused according to the guidelines of the Declaration of Helsinki and approved by the Institutional Review Board of MEDICAL COLLEGE OF WISCONSIN (PRO00022426, 19 September 2014).

